# Trajectories of Neurological Recovery 12 Months after Hospitalization for COVID-19: A Prospective Longitudinal Study

**DOI:** 10.1101/2022.02.08.22270674

**Authors:** Jennifer A. Frontera, Dixon Yang, Chaitanya Medicherla, Samuel Baskharoun, Kristie Bauman, Lena Bell, Dhristie Bhagat, Steven Bondi, Alexander Chervinsky, Levi Dygert, Benjamin Fuchs, Daniel Gratch, Lisena Hasanaj, Jennifer Horng, Joshua Huang, Ruben Jauregui, Yuan Ji, D. Ethan Kahn, Ethan Koch, Jessica Lin, Susan B. Liu, Anlys Olivera, Jonathan Rosenthal, Thomas Snyder, Rebecca S. Stainman, Daniel Talmasov, Betsy Thomas, Eduard Valdes, Ting Zhou, Yingrong Zhu, Ariane Lewis, Aaron S. Lord, Kara Melmed, Sharon B. Meropol, Sujata Thawani, Andrea B. Troxel, Shadi Yaghi, Laura J. Balcer, Thomas Wisniewski, Steven L. Galetta

## Abstract

**Background/Objectives:** Little is known about trajectories of recovery 12-months after hospitalization for severe COVID.

**Methods:** We conducted a prospective, longitudinal cohort study of patients with and without neurological complications during index hospitalization for COVID-19 from March 10, 2020-May 20, 2020. Phone follow-up batteries were performed at 6- and 12-months post-COVID symptom onset. The primary 12-month outcome was the modified Rankin Scale (mRS) comparing patients with or without neurological complications using multivariable ordinal analysis. Secondary outcomes included: activities of daily living (Barthel Index), telephone Montreal Cognitive Assessment (t-MoCA) and Neuro-QoL batteries for anxiety, depression, fatigue and sleep. Changes in outcome scores from 6 to 12-months were compared using non-parametric paired-samples sign test.

**Results:** Twelve-month follow-up was completed in N=242 patients (median age 65, 64% male, 34% intubated during hospitalization) and N=174 completed both 6- and 12-month follow-up. At 12-months 197/227 (87%) had ≥1 abnormal metric: mRS>0 (75%), Barthel<100 (64%), t-MoCA≤18 (50%), high anxiety (7%), depression (4%), fatigue (9%) and poor sleep (10%). 12-month mRS scores did not differ significantly among those with (N=113) or without (N=129) neurological complications during hospitalization after adjusting for age, sex, race, pre-COVID mRS and intubation status (adjusted OR 1.4, 95% CI0.8-2.5), though those with neurological complications had higher fatigue scores (T-score 47 vs 44, P=0.037). Significant improvements in outcome trajectories from 6- to 12-months were observed in t-MoCA scores (56% improved, median difference 1 point, P=0.002), and Neuro-QoL anxiety scores (45% improved, P=0.003). Non-significant improvements occurred in fatigue, sleep and depression scores in 48%, 48% and 38% of patients, respectively. Barthel and mRS scores remained unchanged between 6 and 12-months in >50% of patients.

**Discussion:** At 12-months post-hospitalization for severe COVID, 87% of patients had ongoing abnormalities in functional, cognitive or Neuro-QoL metrics and abnormal cognition persisted in 50% of patients without a prior history of dementia/cognitive abnormality. Only fatigue severity differed significantly between patients with or without neurological complications during index hospitalization. However, significant improvements in cognitive (t-MoCA) and anxiety (Neuro-QoL) scores occurred in 56% and 45% of patients, respectively, between 6- to 12-months. These results may not be generalizable to those with mild/moderate COVID.

## INTRODUCTION

Neurological events in the context of acute SARS-CoV-2 infection have been reported in 12-80% of hospitalized patients^1–3^ and contribute to worse in-hospital^1–3^ and early post-discharge outcomes^4^. In a prospective study of 4,491 consecutive patients hospitalized with COVID, we previously reported a 38% increased hazard of in-hospital death and a decreased likelihood of discharge home among patients diagnosed with a neurological disorder by a board-certified neurologist compared to contemporaneous COVID controls without a neurological event^1^. We then followed patients with neurological events post-discharge, along with a propensity-score matched contemporaneous COVID control group without neurological events hospitalized during the same time frame. We performed 6-month structured follow-up interviews, which included standardized functional, cognitive and neuropsychiatric batteries, and identified persistent deficits in over 90% of patients. Patients with neurological complications during index hospitalization had significantly worse 6-month functional outcomes, more severely impaired activities of daily living, and reduced ability to return to work compared to those without neurological complications^4^. Other larger COVID cohorts have demonstrated similar degrees of disability up to 6-months after discharge^5, 6^, however, there is a paucity of data examining long-term functional and cognitive outcomes or trajectories of recovery. Indeed, studies that have assessed 12-month post-COVID outcomes have focused primarily on subjective symptoms^7, 8^, rather than using objective, standardized outcome batteries. Given that the acute COVID illness is protracted in many patients, measurement of outcomes at 6-months may be too soon to adequately capture the full recovery potential of these patients.

In this prospective, longitudinal study of 12-month outcomes following severe COVID, we aimed to evaluate objective measures of functional status (mRS, Barthel Index of Activities of Daily Living), and cognition (t-MoCA), as well as patient-reported quantified psychiatric metrics (NeuroQoL anxiety, depression, fatigue and sleep). Our primary outcome of interest was a comparison of mRS scores between patients with and without neurological events during index hospitalization. In addition to comparing secondary 12-month outcomes between these groups, we also aimed to assess trajectories of recovery over time in a subset of patients that had testing at both 6- and 12-months post COVID. We hypothesized that those with neurological events would continue to do worse than those without at 12-months, but that there would be an overall improvement across metric scores.

## METHODS

### Study Design and Patient Cohort

We conducted a prospective, observational outcome study of consecutive COVID-19 patients hospitalized at four New York City area hospitals within the same hospital system between March 10, 2020 and May 20, 2020 (Study of Neurologic and Psychiatric Events in Acute COVID-19 [SNaP Acute COVID]. Detailed enrollment, methodology and outcomes at hospital discharge and at 6-months post-COVID onset have been previously reported^1, 4^. Patients were prospectively evaluated by a team of neurologists for development of new neurological disorders during acute COVID-19 hospitalization. Recrudescence of old neurological deficits in the context of acute infection were not counted as new neurological events. A total of 4,491 patients were included in SNaP Acute COVID (N=606 with new neurological events and N=3,885 without new neurological events)^1^. Patients with new neurological disorders who survived to discharge were then propensity score-matched by age, sex and severity of illness (intubation status) to COVID-19 patients *without* neurological events hospitalized during the same period, and 6- and 12-month interviews were conducted. Inclusion criteria were: age ≥18 years, hospital admission, reverse-transcriptase-polymerase-chain-reaction (RT-PCR) positive SARS-CoV-2 infection from nasopharyngeal sampling, survival to discharge and consent to participate in a follow-up interview. Surrogate consent was allowed. Exclusion criteria were: negative or missing SARS-CoV-2 RT-PCR test, or evaluation in an outpatient or emergency department setting only.

### Neurological Diagnoses and Severity of Illness Scales

Neurological diagnoses --including toxic-metabolic encephalopathy, hypoxic-ischemic encephalopathy, stroke (ischemic or hemorrhagic), seizure, neuropathy, myopathy, movement disorder, encephalitis/meningitis, myelopathy, myelitis—followed established criteria ^9–18^. A second review of neurological diagnoses was performed by relevant subspecialty co-authors (e.g. epilepsy, stroke, neurocritical care sub-specialists). Patients could be coded for more than one neurological complication.

Demographic data, past medical/neurological history, clinical course and hospital outcomes (e.g. discharge disposition, hospital length of stay) were collected. Severity of illness during hospitalization was assessed using the Sequential Organ Failure Assessment (SOFA) score^19^ and requirement for intubation. Pre-COVID baseline functional status was assessed with mRS scores as reported by patients and/or their surrogate.

### Study Outcomes

Longitudinal follow-up assessments were conducted by telephone interview among case and control hospital survivors or their surrogates who consented to participate. Contact was attempted at 6-months (± 1 month and 12-months ((± 2 months from the onset of neurological symptoms among cases, or from the onset of COVID-19 symptoms among controls. As previously published, the median time from general COVID-19 symptom onset to neurological complication in this cohort was 2 days^1^. Three attempts at contact were required before patients/surrogates were coded as “unreachable”. We have previously reported outcomes at 6-months^4^, and here present 12-month outcomes and trajectories of metrics over time.

The primary outcome was the mRS [0=no symptoms, 6=dead]^20^, analyzed using ordinal logistic regression. Secondary outcomes included: the Barthel Index^21^ of activities of daily living (0=completely dependent, 100=independent for all activities), the t-MoCA; 22=perfect score; ≤18=abnormal cognition)^22^, and Quality of Life in Neurological Disorders^23^ (NeuroQoL) short form self-reported health measures of anxiety, depression, fatigue and sleep. Patients with fewer than 13 years of education received an additional point when scoring the t-MoCA^24^. NeuroQoL raw scores were converted into T-scores with a mean of 50 and standard deviation of 10 in a reference population (U.S. general population or clinical sample)^25^. Higher T-scores indicate worse self-reported health for the anxiety, depression, fatigue and sleep metrics. NeuroQoL T-scores were considered abnormal if they were >1 standard deviation above the mean (i.e. T-score >60). All of the above batteries have been validated for surrogate completion with the exception of the t-MoCA, which was only scored if the patient was able to complete the assessment. Incomplete or partial responses to a given metric were excluded from analysis. Patients were assessed as being “abnormal on at least one metric” if any of the metrics measured were abnormal (as defined above). If all of the metrics were in normal range, but some scales were not completed, the patient was excluded from coding for this measure.

### Outcome Trajectories

mRS, Barthel and t-MoCA scores were coded as “unchanged” if scores were exactly the same at 6 and 12-months, and worse or improved if scores differed at all. Anxiety, depression, fatigue and sleep NeuroQoL T-scores were coded as worse if there was a ≥1 point increase or improved if there was a ≥1 point decrease in scores between 6- and 12-month measurements.

### Standard Protocol Approvals and Patient Consents

This study was approved by the NYU Grossman School of Medicine Institutional Review Board. All patients or their surrogates provided consent for participation.

### Statistical Analyses

Demographic variables, past medical history, clinical course and in-hospital outcomes were compared between COVID-19 patients with and without a new neurological event using Mann-Whitney U tests for non-normally distributed continuous variables and Chi-square or Fisher’s exact tests for categorical values, as appropriate. For patients who died, a mRS score of 6 was assigned, but no other outcome variables were scored.

Multivariable ordinal logistic regression models predicting 12-month mRS scores were constructed to determine the impact of neurological complications during COVID-19 hospitalization, adjusting for previously identified, clinically relevant demographic and clinical covariates^4^. Secondary outcomes were compared between those with or without neurological disorders using Mann-Whitney U (Wilcoxon rank-sum) or Chi-squared tests as appropriate. Multivariable logistic regression models adjusting for relevant covariates, and respecting the one in ten rule to avoid overfitting the models, were constructed to identify the effect of neurological complications on dichotomized secondary outcomes when univariate analysis yielded a P<0.05. Kaplan-Meier survival analysis was conducted to compare differences in survival distributions between hospital discharge and 12-month follow-up among those with and without neurological events. Dates of death were collected by chart review and surrogate interview.

Next, among a subgroup of patients who completed *both* 6-month and 12-month follow-up interviews, non-parametric paired samples sign tests were used to compare continuous, non-normally distributed, non-symmetrical metric scores over time. Similarly, nonparametric related-samples McNemar tests were used to compare dichotomous outcomes over time. Correlations between different outcome measures were assessed using Spearman correlation coefficients. All analyses were conducted using IBM SPSS Statistics for Mac version 25 (IBM Corp., Armonk, NY).

## RESULTS

### Enrollment Characteristics

Of 715 patients eligible for a 12-month follow-up, contact was attempted with 590 patients. 242/590 (41%) completed the 12-month follow-up interview (Figure 1). Calls were not attempted in 125 patients due to language barrier, missing or defunct contact information or indication at the 6-month call that the respondent did not wish to be re-contacted. Of the 348 calls that were attempted but not completed, N=270 were unreachable after 3 phone calls, N=7 were unable to understand the consent process and N=67 refused participation. In the entire cohort, the median age was 65 years (IQR 53-73) and 155/241 (64%) were male. Of the 242 included patients, N=113 (47%) had neurological complications during index hospitalization and N=129 (53%) had no neurological complications during hospitalization. The median time from neurological symptom onset following SARS-CoV-2 infection (or COVID-19 symptom onset for controls) to follow-up interview was 393 days (IQR 375-409 days). Among those able to participate in outcome assessments, direct patient responses were obtained from 84/113 (74%) neurological patients and 110/129 (85%) controls, while surrogate responses were utilized in 29/113 (26%) neurological patients and 19/129 (15%) controls (P=0.199). Because some interviews were completed by surrogates, and some patients were too impaired to participate in cognitive or NeuroQoL testing, not all metrics were completed by all patients: 235/242 (97%) completed mRS scoring, 236/242 (98%) completed the Barthel, 170/242 (70%) completed the t-MoCA, 223/242 (92%) completed the fatigue NeuroQoL, 221/242 (91%) completed the sleep NeuroQoL, 225/242 (93%) completed the anxiety NeuroQoL and 225/242 (93%) completed the depression NeuroQoL.

**Figure 1.**
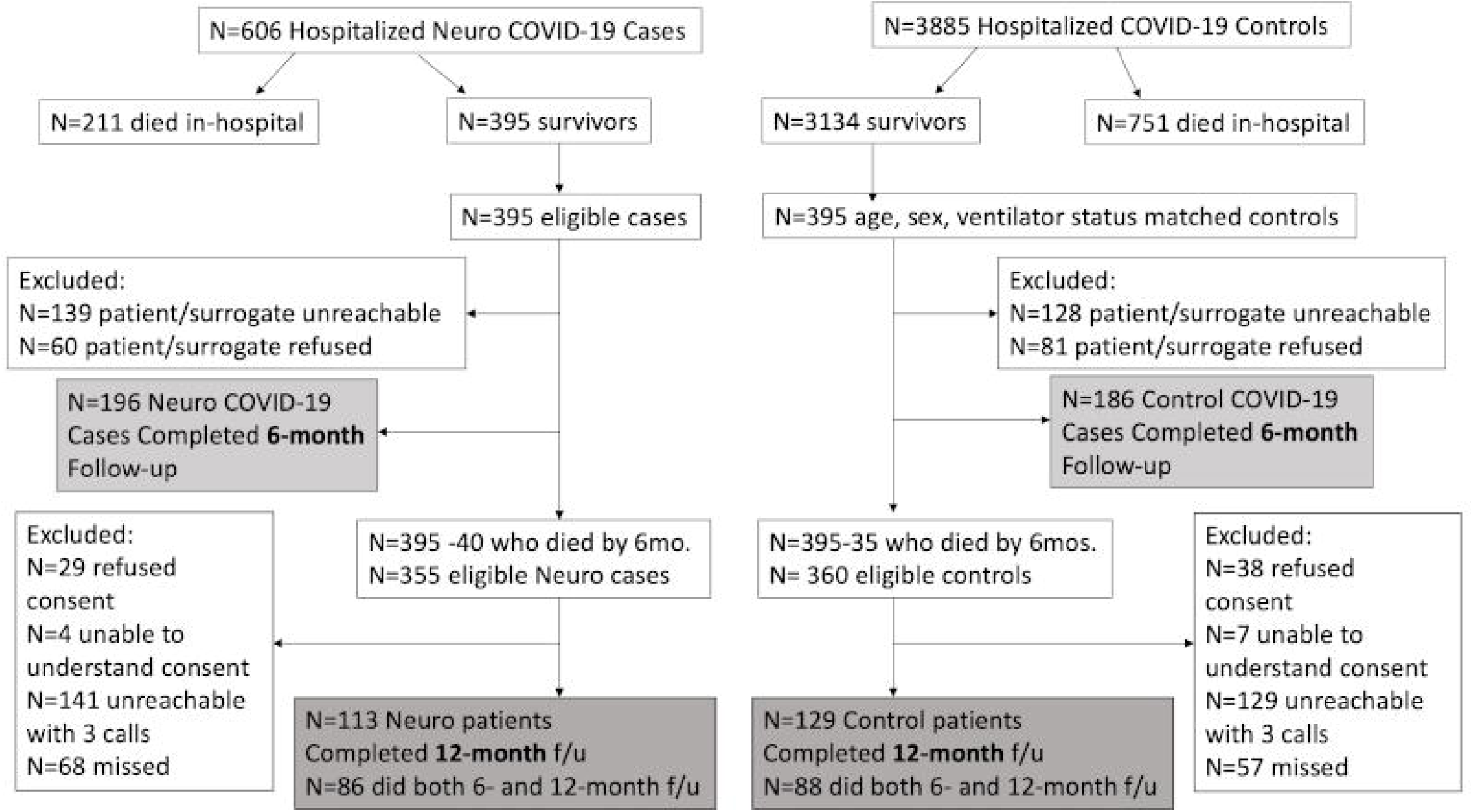
Study Enrollment: Enrollment flow chart identifying inclusion and exclusion of patients with or without neurological complications from index COVID-19 hospitalization to 6-month and 12-month follow-up. N=395 COVID-19 patients with neurological complications who survived index hospitalization were propensity score matched by age, sex and intubation status to N=395 COVID-19 patients hospitalized during the same time frame and without neurological complications, who survived to discharge.

### Demographic and Clinical Differences between Groups

At 12-months, compared to controls without neurological complications, those with neurological events during hospitalization more often had a past history of seizure or dementia, but were otherwise similar in age, sex, race/ethnicity, and medical comorbidity rates (Table 1). Most neurological events occurred in patients without a prior history of similar events. For example, none of the patients diagnosed with ischemic or hemorrhagic stroke had a prior history of stroke, and only 1 of 10 patients with a newly diagnosed movement disorder, had a prior movement disorder history. However, 6 of 12 patients who developed seizure in the context of acute COVID had a prior seizure disorder. Markers of severity of COVID-19 illness during index hospitalization, including intubation (33% in neurological complication patients versus 34% in controls, P=0.822), SOFA scores, minimum oxygen saturation and minimum blood pressure, did not differ significantly between groups. Rate of discharge to a nursing home was slightly higher among patients with neurological events, while more patients were discharged to long term acute care hospitals in the control group.

**Table 1.**
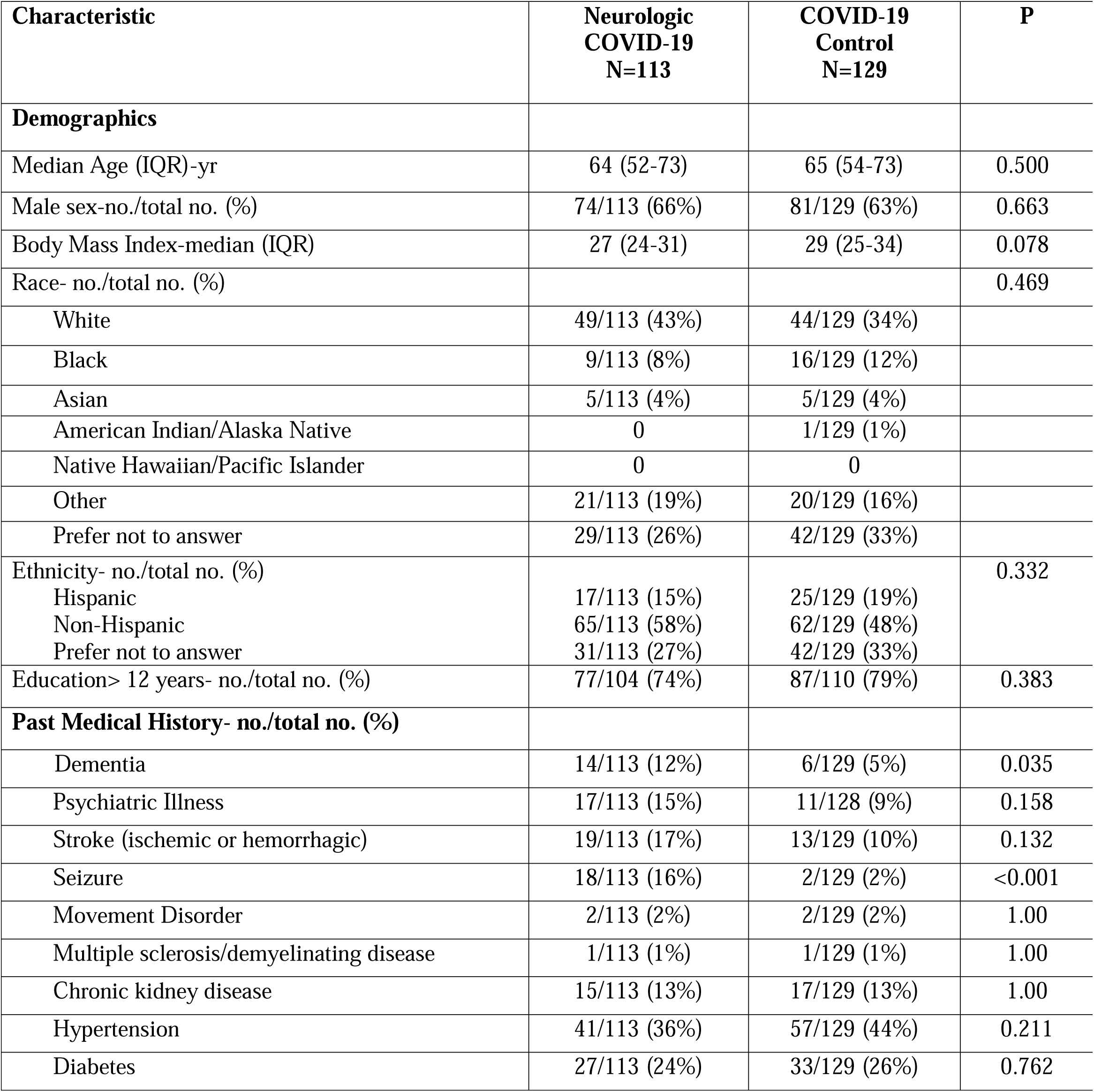
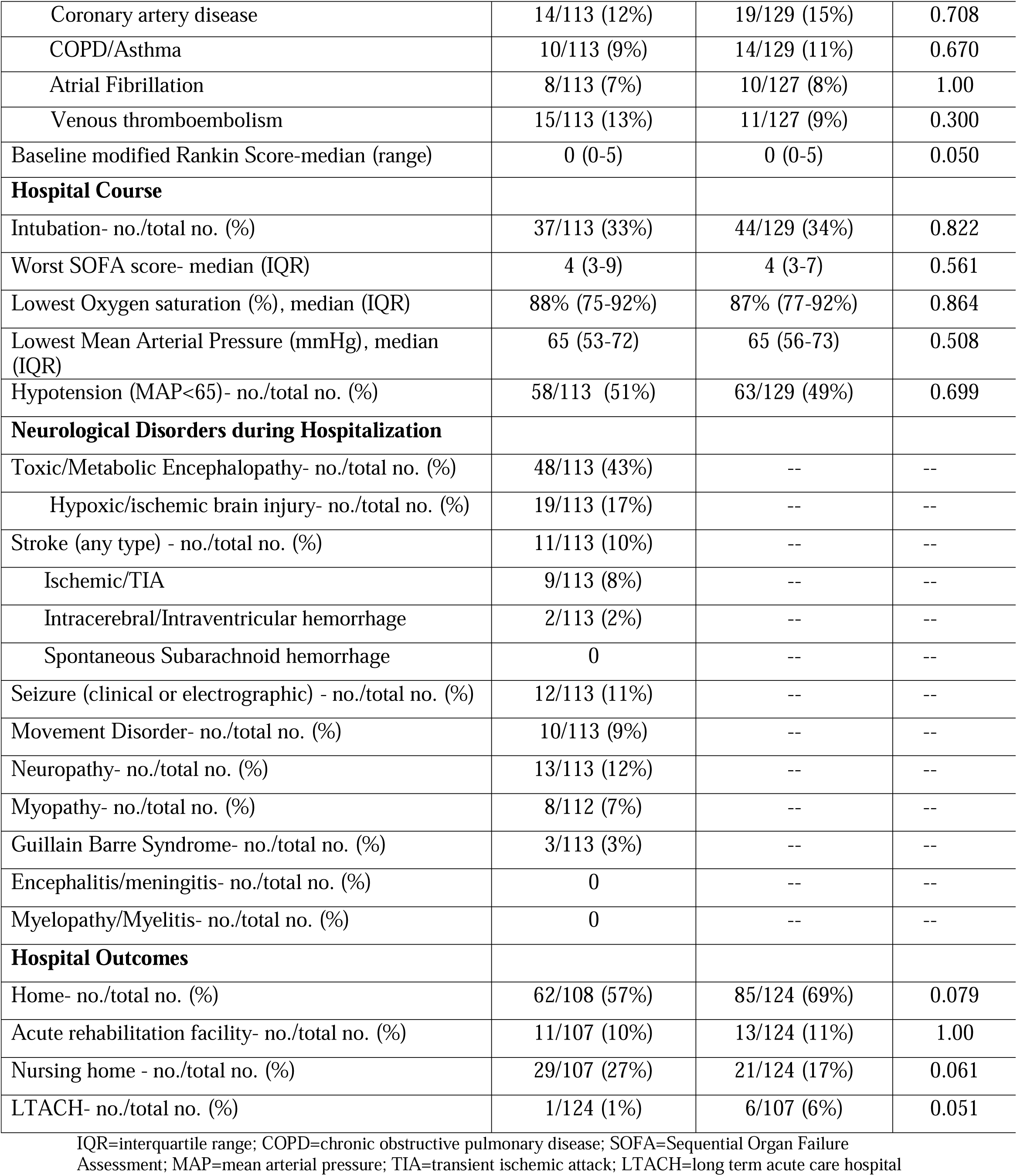
Demographics, hospital course and discharge disposition among patients with and without neurological events during index COVID-19 hospitalization

### Primary Outcome: Modified Rankin Scale

At 12-months, 26/110 (24%) of patients with neurological complications had no disability or mild symptoms that didn’t interfere with activities (mRS 0-1) compared to 49/125 (39%) of patients without neurological complications (P=0.011). There was a suggestion of worse mRS scores in those with neurological complications (median mRS 3 [IQR 2-4]) compared to controls (median mRS 2 [IQR 0-4]; univariate ordinal logistic regression odds ratio 1.54, 95% CI 0.98-2.43, Wald X^2^(1)=3.46, P=0.063). However, in multivariable ordinal logistic regression analysis after adjusting for age, sex, race, pre-COVID mRS and intubation status, there was no significant difference in 12-month mRS scores between patients with or without neurological complications during index hospitalization (adjusted odds ratio for worse mRS scores 1.4, 95% confidence interval 0.8-2.5, P=0.258, Figure 2). Additional sensitivity analyses using models adjusting for past neurological history did not change these results.

**Figure 2.**
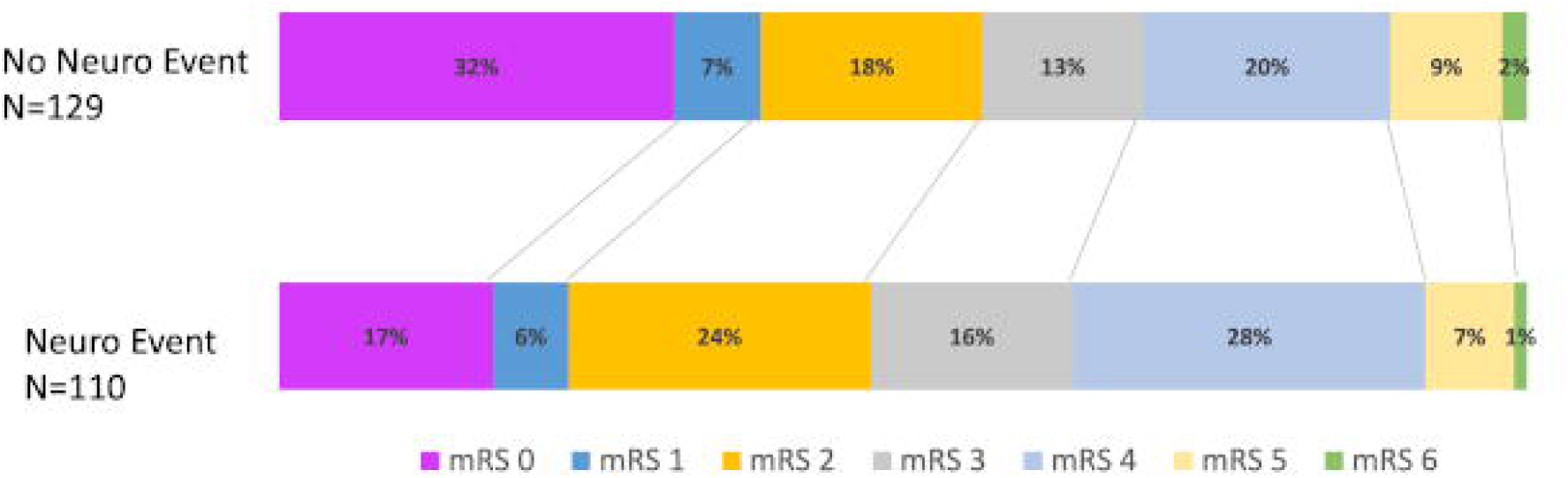
Ordinal analysis of 12-month Modified Rankin Scale (mRS) scores: MRS scores 12-months post COVID-19 symptom onset in patients with and without neurological complications during index COVID-19 hospitalization. There was no significant difference between groups in mRS scores using multivariable ordinal logistic regression analysis adjusting for age, sex, race, pre-COVID mRS and intubation status (adjusted odds ratio for worse mRS scores 1.4, 95% confidence interval 0.8-2.5, P=0.258). mRS 0=no symptoms or disability, 1=symptoms, but no disability, 2=slight disability, unable to carry out all previous activities, but able to function without assistance, 3=moderate disability, requiring help, but able to walk without assistance, 4=moderately severe disability, unable to walk or attend to bodily needs without assistance, 5=severe disability, bedbound, incontinent, requiring constant care, 6=dead.

Between 6-months and 12-months, 1/113 (1%) patients with neurological events and 2/129 (2%) of controls died (P=1.00). A total of 451 patients completed 6-month and/or 12-month follow-up interviews. Of these, 78/451 (17%) died between hospital admission and 6 or 12-month follow-up (N=41/220 [19%] with neurological events and N=37/231 [16%] without neurological events, Kaplan-Meier log rank P=0.379, e Figure 1 in the Supplement).

### Secondary Outcomes

Overall, 197/227 (87%) of patients who completed all 12-month follow-up batteries had at least one abnormal metric (Table 2): 176/235 (75%) had a mRS>0, 150/236 (64%) had a Barthel Index of Activities of Daily Living<100, and 80/161 (50%) of patients without a prior history of dementia/cognitive abnormalities had an abnormal T-MoCA score (≤18, e Table 1 in the Supplement). NeuroQoL scores > 1 standard deviation above the mean occurred in 16/225 (7%) for anxiety scores, 9/225 (4%) for depression scores, 20/223 (9%) for fatigue scores and 22/221 (10%) for sleep scores. Excluding patients with a pre-COVID mRS>0, 107/130 (82%) of patients had at least one abnormal metric at 12-months.

**Table 2.** Secondary outcomes at 12-months post COVID-19 symptom onset compared between patients with or without neurological events during index COVID-19 hospitalization

|  | **Neurologic COVID-19**<br>**N=113** |  | **COVID-19 Control**<br>**N=129** |  |  |
| --- | --- | --- | --- | --- | --- |
|  | Total N | Median, IQR<br>or N (%) | Total N | Median, IQR<br>or N (%) | P |
| **Activities of Daily Living** |  |  |  |  |  |
| Barthel Index | 111 | 100 (75-100) | 125 | 100 (90-100) | 0.282 |
| Any abnormal activities of daily living (Barthel<100) | 111 | 68/111 (61%) | 125 | 82/125 (66%) | 0.489 |
| **Cognitive Outcomes** |  |  |  |  |  |
| Telephone MOCA | 76 | 19 (15-20) | 94 | 18 (15-20) | 0.931 |
| Abnormal t-MOCA ($\leq 18$) | 76 | 37/76 (49%) | 94 | 48/94 (51%) | 0.758 |
| Abnormal t-MOCA ($\leq 18$)<br>excluding h/o dementia | 69 | 33/69 (48%) | 92 | 47/92 (51%) | 0.682 |
| **Neuro Quality of Life** |  |  |  |  |  |
| Anxiety T-score | 105 | 47 (39-53) | 120 | 44 (36-53) | 0.301 |
| Depression T-score | 105 | 43 (37-50) | 120 | 43 (37-49) | 0.480 |
| Fatigue T-score | 105 | 47 (38-54) | 118 | 44 (37-51) | 0.035 |
| Sleep T-score | 103 | 46 (39-54) | 118 | 44 (36-53) | 0.247 |
| Worse than average Anxiety, (T-score>60) | 105 | 9/105 (9%) | 120 | 7/120 (6%) | 0.448 |
| Worse than average Depression, (T-score>60) | 105 | 7/105 (7%) | 120 | 2/120 (2%) | 0.086 |
| Worse than average Fatigue, (T-score>60) | 105 | 15/105 (14%) | 118 | 5/118 (4%) | 0.010 |
| Worse than average Sleep, (T-score>60) | 103 | 9/103 (9%) | 118 | 13/118 (11%) | 0.656 |
| Total patients with at least one abnormal metric | 108<br>56* | 99/108 (92%)<br>49/56 (88%)* | 119<br>74* | 98/119 (82%)<br>58/74 (78%)* | 0.039<br>0.177* |

t-MoCA=telephone Montreal Cognitive Assessment; IQR=interquartile range. Bold indicates P<0.05
\*Excluding pts with mRS>0 baseline.

There were significant correlations between t-MoCA scores and depression T-scores (Spearman’s rho −0.187, P=0.016), anxiety and depression T-scores (Spearman’s rho 0.716, P<0.001), anxiety and fatigue T-scores (Spearman’s rho 0.674, P<0.001), anxiety and sleep T-scores (Spearman’s rho 0.655, P<0.001), depression and fatigue T-scores (Spearman’s rho 0.596, P<0.001), depression and sleep T-scores (Spearman’s rho 0.624, P<0.001) and sleep and fatigue T-scores (Spearman’s rho 0.736, P<0.001).

Comparing those with versus without neurological complications, only fatigue scores differed significantly in univariate analysis (14% of neurological patients had T-scores>60 compared to 4% of patients without neurological complications, P=0.010). In multivariable logistic regression analysis adjusting for age, ventilator status and baseline mRS, patients with neurological complications more often had severe fatigue than controls (adjusted OR 3.18, 95% CI 1.1-9.4, P=0.037). Of the 15 patients with neurological deficits and severe fatigue, 7/15 (47%) had toxic-metabolic encephalopathy, 4/15 (27%) had seizure, 2/15 (13%) had movement disorders, 1/15 (7%) had Guillain-Barre Syndrome, and 1/15 (7%) had neuropathy. More patients with neurological events had at least one abnormal metric (92%) compared to controls (82%, P=0.039).

### Outcome Trajectories from 6 to 12-months post COVID onset

Next, we evaluated changes in outcome metrics over time. Overall, 174 patients completed both 6- and 12-month follow-up interview (N=86 with neurological events and N=88 controls). Raw scores for patients who completed functional, cognitive and NeuroQoL metrics at *both* 6- and 12-months are reported in Table 3, and scores for all patients at each time frame are shown in e Table 1 in the Supplement. A 6-months, 150/171 (88%) had at least one abnormal metric compared to 141/168 (84%) patients at 12-months (paired-samples McNemar test P=0.230). When excluding patients with pre-COVID baseline mRS>0, results were similar (83% had at least one abnormal metric at 6-months versus 80% at 12-months, McNemar test P=0.503).

**Table 3.** Functional, cognitive and patient-reported outcome metrics among patients that completed both 6-months and 12-months post-COVID interviews (N=174)

| **Outcome Measure** | **Baseline** | **6-month**<br>**N=174** | **12-month**<br>**N=174** |
| --- | --- | --- | --- |
| mRS<br>median (IQR) | 0 (0-1) | 3 (1-4) | 3 (0-4) |
| mRS>0, N (%) | 54/171 (32%) | 310/380 (82%)<br>82/116 (71%)* | 127/171 (74%)<br>77/115 (67%)* |
| Barthel<br>median (IQR),<br>Barthel <100, N (%) | --<br>-- | 100 (75-100)<br>76/171 (44%) | 100 (85-100)<br>64/170 (38%) |
| tMoCA<br>median (IQR) | -- | 18 (15-20) | 19 (15-21) |
| tMOCA ≤18, N (%) | -- | 70/126 (56%) | 57/127 (45%) |
| tMOCA (≤18) excluding h/o<br>dementia, N (%) | -- | 62/117 (53%) | 52/119 (44%) |
| Neuro-QoL Anxiety T-score,<br>median (IQR) | -- | 50 (42-56) | 47 (36-53) |
| Anxiety T-score>60, N (%) | -- | 13/163 (8%) | 9/161 (6%) |
| Neuro-QoL Depression T-score,<br>median (IQR) | -- | 44 (37-51) | 43 (37-50) |
| Depression T-score>60, N (%) | -- | 7/162 (4%) | 8/161 (5%) |
| Neuro-QoL Fatigue T-score,<br>median (IQR) | -- | 47 (40-53) | 46 (38-53) |
| Fatigue T-score>60, N (%) | -- | 9/158 (6%) | 15/159 (9%) |
| Neuro-QoL Sleep T-Score,<br>median (IQR) | -- | 47 (39-54) | 46 (36-53) |
| Sleep T-score>60, N (%) | -- | 17/162 (11%) | 15/159 (9%) |
| Total patients with at least one<br>abnormal metric | -- | 150/171 (88%)<br>95/114 (83%)* | 141/168 (84%)<br>89/111 (80%)* |

\*Excluding pts with mRS>0 baseline. This percentage indicates the patients with new functional disability after COVID-19 hospitalization
mRS=modified Rankin Scale; t-MoCA=telephone Montreal Cognitive Assessment; IQR=interquartile range; QoL=quality of life

A nonparametric, paired-sample sign test with continuity correction was conducted to evaluate changes in outcome metrics from 6-month to 12-month follow-up. There was a statistically significant median improvement in t-MoCA scores (+1 point) from 6-months (median score 18) to 12-months (median score 19), z= −2.437, P=0.002; and a statistically significant improvement in NeuroQoL anxiety scores from 6-months (median score 49.5) to 12-months (median score 47.3), z=−3.164, P=0.002. Overall, 56% of patients had improved t-MoCA scores over time and 45% had improved anxiety scores (Figure 3). While there were no statistically significant changes, improvements were seen in 48% of fatigue scores, 48% of sleep scores and 38% of depression scores. Conversely, the majority of patients showed no changes in mRS or Barthel scores between 6- and 12-month follow-up.

**Figure 3.**
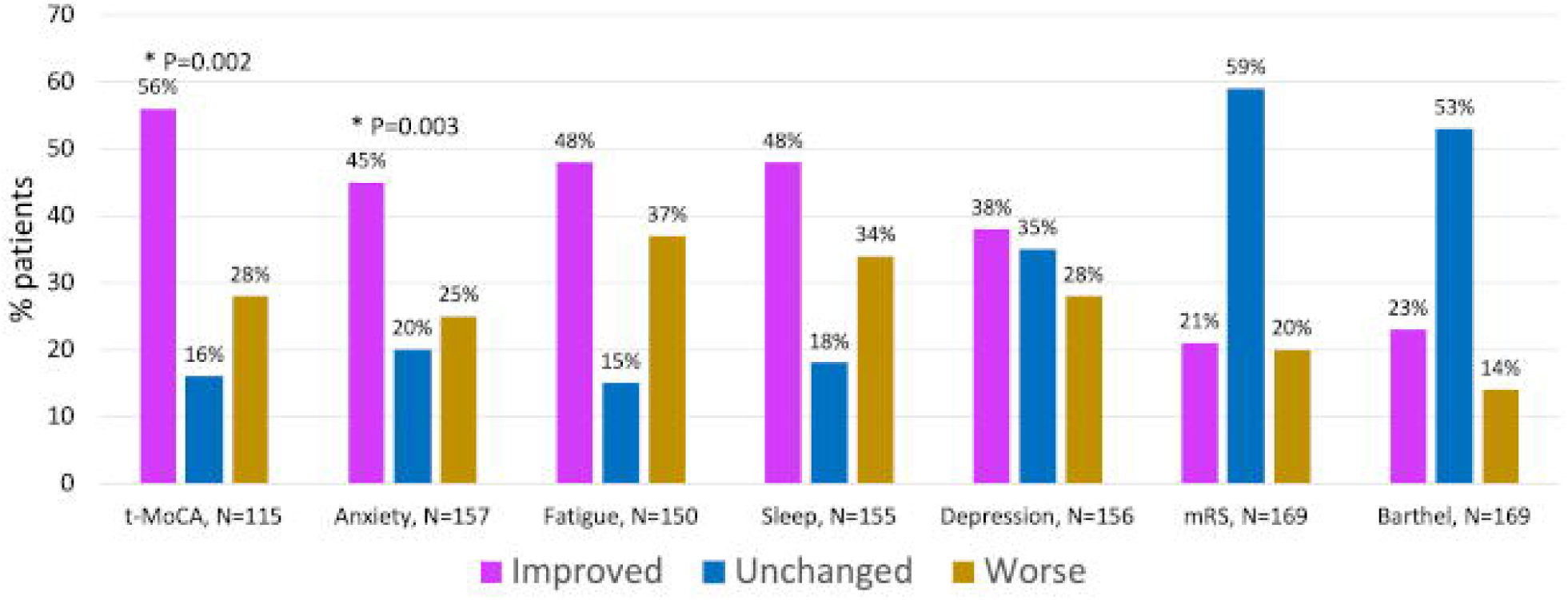
Percent of patients with improved, worse or same outcome scores between 6- and 12-months post COVID Hospitalization (N=174)

## DISCUSSION

In this prospective cohort study, we found that at 12-months post-severe COVID onset, fewer than 20% of all patients scored within the normal range on all of the assessed metrics. Patients with neurological complications fared worse, as only 8% of neurological patients scored in the normal range on all metrics, compared to 18% of control patients. Although we did not find a significant impact of neurological events during index COVID hospitalization on the primary mRS outcome at 12-months, there was a greater than 3-fold significantly increased odds of severe fatigue among neurological patients, even after adjusting for confounders. Additionally, abnormal scores on cognitive testing persisted in 50% of patients without a pre-COVID history of cognitive abnormalities, irrespective of the presence or absence of a neurological complication during hospitalization. Indeed, rates of abnormal cognition were substantially higher than rates of abnormalities in other domains such as activities of daily living, anxiety, depression, fatigue or sleep, indicating that cognition should be an area of focus for post-acute COVID study.

When examining trajectories of recovery from 6 to 12-months, the majority of patients did not have improvements in functional status (mRS) or activities of daily living (Barthel Index), however, there were significant improvements in cognition and anxiety scores between 6-month and 12-month follow-up, and substantial proportions of patients had non-significant improvements in fatigue, sleep and depression scores. While some studies have reported that 45%-77% of patients have at least one symptom 12-months after COVID hospitalization^7, 8, 26^, we identified higher rates of objective abnormalities in functional, cognitive and self-reported patient-centered outcomes. In contrast to subjective symptom-based data, our findings highlight that standardized metrics may unmask disability or psychiatric symptoms of which patients may not even be cognizant. A smaller longitudinal study of 51 patients assessed at 12-months post-COVID ARDS (adult respiratory distress syndrome) utilized standardized batteries and identified severe fatigue in 26% of patients and abnormal 6-minute walk test in 36% of patients^26^, though only 7/51 (16%) of patients had an abnormal MoCA score (<25). Because this study included only patients who were capable of in person follow-up, the most severely affected patients were likely not captured. Additionally, although metrics were assessed at 3, 6 and 12-months post-COVID, the authors were only able to report aggregate findings, rather than assess improvements over time since, few patients had more than one visit. To our knowledge, our study represents the largest prospective cohort study to report functional, cognitive and psychiatric metrics in patients with severe COVID at 12-months post infection, and the first to report repeated assessments and trajectories of recovery over time. Other strengths of our study include its prospective ascertainment of in-hospital new neurological complications diagnosed by neurologists, and multi-domain, long-term serial follow-up structure.

While we previously reported higher in-hospital mortality rates^1^ and worse 6-month mRS scores^4^ for patients with neurological complications compared to contemporaneous COVID controls without neurological events, the difference in mRS scores between groups was no longer significant at 12-months in multivariable analysis. This finding may be attributable to neurological improvement over time, or the smaller number of patients that completed 12-month follow-up, which would mean less power to detect small differences. Because there were fewer neurological patients who completed the 12-month interview than controls, it’s possible that some of the most debilitated neurological patients were unable to complete follow-up or their surrogates were unwilling to participate. Since a higher proportion of patients with neurological complications died in-hospital compared to those without^1^, our findings represent a more mild spectrum of sequelae following neurological events in COVID, and we may be underestimating the impact of neurological injury.

The finding of higher rates of severe fatigue among COVID patients with neurological events is not entirely surprising given the high rates of chronic fatigue observed in a variety of neurological disorders including stroke, multiple sclerosis, dysautonomia, traumatic brain injury and neuromuscular disorders^27^. The underlying pathophysiology of fatigue may be related to diencephalic, hypothalamic/pituitary lesions, neurotransmitter imbalances or interruption of cortical and basal ganglia circuits^27^. Pro-inflammatory cytokines have also been implicated in the pathogenesis of fatigue^28^ and COVID-related hyperinflammation coupled with concomitant neurological injury may exacerbate fatigue symptoms. Interestingly, we did not observe differences in sleep metrics between the two groups, though sleep and fatigue T-scores were very highly correlated among individuals.

Improvements in t-MoCA scores over time should generate a degree of optimism regarding recovery from long-COVID, particularly since “brain fog”, confusion and dysexecutive function appear to be common protracted post-acute sequelae^29–31^. While the pathogenesis of post-acute cognitive dysfunction is likely multifactorial, possibilities include post-hypoxic brain injury, blood brain barrier disruption with ongoing inflammation, autoimmune mechanisms or even neurodegenerative disease^32^. Indeed, significant decay in pre-morbid to post-COVID MoCA scores has been documented in over 20% of patients with only mild symptomatic COVID. In this study, COVID patients were found to have an 18-fold increased odds of developing cognitive decline compared to seronegative controls^33^. Though we found a median of only 1 point improvement in t-MoCA scores over a 6-month interval (between 6- and 12-month follow-up), this likely represents a clinically significant change based on data from the 30-item standard MoCA, which indicates that a 1.7 point change over a much longer 3.5 year follow-up represents a clinically meaningful difference^34^.

Similarly, post-COVID psychiatric sequelae-particularly anxiety disorders-are common and occur at a higher frequency than has been observed in individuals with influenza or other respiratory tract infections^35^. The improvements we observed in NeuroQoL anxiety scores were smaller, yet still significant and may be related to improvements in or acclimation to pandemic related stressors, therapeutic intervention or biological recovery. There is likely an interplay between psychiatric well-being and cognitive function, as evidenced by the significant correlations we observed between t-MoCA scores and depression T-scores. However, we did not detect significant improvements in depression scores over time, which implies that improvements in depression are not likely the driving factor for cognitive improvements.

There are limitations to this study. First, it is possible that we observed a practice effect with the t-MoCA, rather than a true cognitive improvement, however, test-retest reliability studies evaluating the full 30-item MoCA at a shorter time frame (1 month) demonstrated a <1 point mean change in scores^36^. Additionally, the t-MoCA is a screening test, is not as sensitive as formal neuropsychometric testing, and may be biased by race or social determinants of health^37, 38^, though we did adjust scores for education level. Although we did a sensitivity analysis excluding patients with a pre-COVID history of dementia or cognitive impairment, we did not have pre-COVID t-MoCA scores to assess the degree of change following COVID hospitalization. Future studies utilizing longitudinal cohorts with pre-COVID data may be able to more precisely assess the cognitive impact of severe COVID. Second, because we allowed for surrogate responses not all metrics were completed on all patients, which may have limited our power to detect small differences. Additionally, outcomes may be worse than we estimated as the sickest patients were not able to participate in some of the testing and family members of patients who did poorly may have been less motivated to participate in research. Third, we did not have a SARS-CoV2 negative control group. It’s possible that pandemic-related societal, economic and environmental stressors may have contributed to certain outcomes (notably psychiatric sequelae) rather than being biologically driven by SARS-CoV-2. Last, we may not have captured all neurological complications during hospitalization, particularly among the sickest patients who could not tolerate being off sedation for assessment. Hence, we may have underestimated the impact of neurological complications on long-term outcomes.

## CONCLUSIONS

At 12-months post-severe COVID, >80% of patients who had no baseline functional abnormalities (mRS 0), had at least one abnormal measure of functional, cognitive or neuropsychiatric outcome. Notably, abnormal cognitive testing occurred in 50% of patients who did not have a pre-COVID history of dementia or cognitive abnormalities. We did not detect an impact of neurological events during index COVID-hospitalization on the primary outcome of mRS scores, though patients with neurological complications had significantly higher rates of severe fatigue compared to controls. Among the subset of patients who underwent serial assessments at 6- and 12-months post-COVID, there were significant improvements in cognition and anxiety scores. Given the high morbidity burden at 1-year following COVID onset, research focusing on therapeutic interventions is warranted.

## Data Availability

All data produced in the present study are available upon reasonable request to the authors

## ACKNOWLEDGEMENTS/CONFLICTS/FUNDING

We would like to thank the patients and families who participated in this study. There are no direct funding sources to report for this study.

JAF receives/received funding for the following COVID-related grants: NIH/NINDS 3U24NS11384401S1, NIH/NHLBI 1OT2HL161847-01, NIH/NIA 3P30AG066512-01; ABT receives funding for the following COVID-related grants: NIH/NINDS 3U24NS11384401S1, NIH/NHLBI 1OT2HL161847-01; SM and SY receive funding for the following COVID-related grant: NIH/NINDS 3U24NS11384401S1; ST, LH and LJB receive funding for the following COVID-related grant: NIH/NHLBI 1OT2HL161847-01; TW received funding for the following COVID-related grant: NIH/NIA 3P30AG066512-01.

## FIGURE LEGENDS

**e Figure 1**. Kaplan-Meier curve of time to death from hospital admission through 12-month follow-up among patients with (N=220) or without (N=231) neurological complications during hospitalization. Patients with 6-month and/or 12-month follow-up interviews were included (total N=451).

